# The Adaptive Olfactory Measure of Threshold (ArOMa-T): A rapid test of olfactory function

**DOI:** 10.1101/2022.03.08.22272086

**Authors:** Elisabeth M. Weir, Mackenzie E. Hannum, Danielle R. Reed, Paule V. Joseph, Steven D. Munger, John E. Hayes, Richard C. Gerkin

## Abstract

Many widely-used psychophysical olfactory tests have limitations that can create barriers to adoption outside research settings. For example, tests that measure the ability to identify odors may confound sensory performance with memory recall, verbal ability, and past experience with the odor. Conversely, typical threshold-based tests avoid these issues, but are labor intensive. Additionally, many commercially-available olfactory tests are slow and may require a trained administrator, making them impractical for use in a short wellness visit or other broad clinical assessment. We tested the performance of the Adaptive Olfactory Measure of Threshold (ArOMa-T) – a novel odor detection threshold test that employs an adaptive Bayesian algorithm paired with a disposable odorant – delivery card – in a non-clinical sample of individuals (n=534) at the 2021 Twins Day Festival in Twinsburg, OH. Participants successfully completed the test in under 3 min with a false alarm rate of 9.6% and a test-retest reliability of 0.61. Odor detection thresholds differed by sex (∼3.2-fold lower for females) and age (∼8.7-fold lower for the youngest versus the oldest age group), consistent with prior studies. In an exploratory analysis, we failed to observe evidence of detection threshold differences between participants who reported a history of COVID-19 and matched controls who did not. We also found evidence for broad-sense heritability of odor detection thresholds. Together, these data indicate the ArOMa-T can determine odor detection thresholds. The ArOMa-T may be particularly valuable in clinical or field settings where rapid and portable assessment of olfactory function is needed.

## 1. Introduction

Olfactory dysfunctions are prevalent and underdiagnosed medical conditions that can have serious consequences for health, diet, safety and quality of life (1, 2). Some consequences include altered diet, a decreased ability to detect dangers such as fire or spoiled food, and feelings of disconnection from the environment and other people. Some estimates suggest up to 1 in 4 people may have some type of olfactory disorder (3). The most common causes for olfactory disorders are head trauma, sinonasal disease, and upper respiratory infections (4, 5), and olfactory disorders are associated with aging (6). Olfactory dysfunction can also be an early biomarker of neurodegenerative diseases, including Alzheimer’s disease and Parkinson’s disease, where olfactory deficits precede detectable memory loss (7). Despite the impact of olfactory disorders on patients and their known associations with other serious health conditions, olfactory function is infrequently tested in routine clinical practice (8).

While the importance of olfactory disorders has historically been neglected despite their prevalence and impact, public awareness of anosmia increased dramatically when sudden smell loss was highlighted as a highly predictive symptom of COVID-19 (e.g. (9)). One meta-analysis suggested ∼75% of COVID-19-positive individuals experienced at least a transient loss of smell (10), while long term smell loss may persist in millions of individuals (11), with substantially negative impact on quality of life (12).

Psychophysical testing employed in clinical settings is typically used to determine if a patient has a quantitative olfactory disorder: either anosmia (a complete or near complete loss of smell) or hyposmia (where the patient’s ability to detect or perceive odors is substantially reduced but not absent). Notably, quantitative tests are not optimized to assess qualitative disorders like parosmia (distorted smell) or phantosmia (distorted smell), which depend on patient report. Quantitative tests typically measure one or more specific parameters: odor identification (“what is this? vanilla!”), odor discrimination (“is this smell different from the last one?”), and odor detection threshold (“what is the lowest concentration the patient can smell?”). The two most common clinical tests are the University of Pennsylvania Smell Identification Test (UPSIT), which is composed of 40 odor identification questions (13), and Sniffin’ Sticks, which measures odor identification, odor discrimination, and odor detection threshold (14) to create a composite score or index of function. In clinical samples, multiple measures of olfactory performance are typically highly, though not fully, correlated (e.g., (3, 15, 16)). Therefore, using just one measure often suffices in normal practice.

Widely used psychophysical tests for diagnosis of hyposmia and anosmia have some limitations. For example, odor identification tasks require the patient to a) smell the stimulus, b) recognize the stimulus from prior experience, and c) communicate the correct name. Thus, this measure confounds sensory performance with memory recall and verbal ability. Odor identification may also be challenging if stimuli are culturally or experientially dependent (e.g., root beer is widely known in the United States, but not Europe or Asia). Thus, odor identification tests should be validated (17) in different populations and global locations prior to mainstream clinical use to obtain appropriate normative data. Further, odor identification tests make cognitive demands that may pose issues for elderly patients or other special populations (e.g., children, or those with cognitive impairments). Odor discrimination tests also have cognitive demands, particularly regarding working memory, that can limit their use in such populations.

By contrast, olfactory tests that measure odor detection threshold have several potential advantages. Such tests are semantics-free, thus avoiding issues of familiarity, naming, and recall (18). They also may be more sensitive and/or specific measures of hyposmia and anosmia. That is, the odorants given in an odor identification task are normally presented at a concentration well above threshold, so a small but real drop in olfactory function may be missed (19). In this case, the drop may not impair the ability of the patient to successfully identify and name the odor, despite the presence of a true quantitative loss. Still, odor detection threshold tests are used much less frequently in clinical settings, in part due to difficulties with stimulus control and test duration. Indeed, threshold estimation using traditional methods (such as a 2-down / 1-up staircase) (20) can take 30 minutes or more to get a single measure of threshold.

Recently, we have developed a novel odor detection threshold test, the Adaptive Olfactory Measure of Threshold (ArOMa-T), which we describe here. This card-based tool is paired with an adaptive Bayesian algorithm, delivered via a simple app, to rapidly guide users through a task that delivers only those stimulus concentrations that will be maximally informative in determining an estimate of the odor detection threshold for that individual. To test the performance of this novel test, we deployed the ArOMa-T among a group of individuals without active COVID-19 who were attending the two-day 2021 Twins Day Festival in Twinsburg, OH.

## 2. Methods

### Participants

The protocol was approved by the local Institutional Review Board for the Monell Chemical Senses Center (*IRB protocol#: 843798*), and the study followed the principles of the Declaration of Helsinki. Per University of Florida requirements, an additional protocol was approved to receive and analyze anonymized aggregate data (*IRB protocol #: IRB202102968*). Participants were recruited, consented, and enrolled at a tent managed by the Monell Center in Twinsburg, OH; 595 participants enrolled in the study between August 7th and August 9th, 2021. All participants provided informed consent electronically. Demographic characteristics of participants (n=534; 29.8% male and 70.2% female; mean age 39.3) are summarized in Table 1. The cohort was predominantly female and white, with a mean age of 39.2 years (± standard deviation of 15.7 years) and median age of 34.1.

**Table 1:** Participant demographics for ArOMa-T Twins Day study after data cleaning

*Participants*
|  |  |
| --- | --- |
| **Sex** |  |
| Male | 159 |
| Female | 375 |
| **Race/Ethnicity** |  |
| White | 459 |
| Black/African American | 35 |
| Amer. Indian/Alaska Native | 5 |
| Hispanic/Latino | 4 |
| Other | 6 |
| Prefer not to answer | 5 |
| Did not answer | 20 |
| **Age (mean ± SD) in years** | 39.2 ± 15.7 |

Some participants were excluded from the analyses (Fig. 1). Participants (n=35) that had an indeterminate threshold from inconsistent response patterns were excluded. Because of the very small number of participants who did not indicate sex as male or female (n = 4), these individuals were excluded from the analysis. The participant pool was highly age diverse, so we classified participants into three age bins of equal size (in years) for analyses: young (18-37 years), middle aged (38-57 years), and older (58-77). Participants (n=22) who were younger than 18 years or older than 78 years were excluded from the analysis (to facilitate the construction of equally-sized age bins). This resulted in a final dataset of 534 unique individuals (Fig. 1); of these, 78 participants (∼15%) reported a prior case of COVID-19. For participants reporting positive COVID-19 history, no data on elapsed time between testing and COVID-19 were collected, so we cannot speak to speed of recovery. Also, a subset of participants (n=97) at the Twins Day Festival returned the next day to repeat the test; these data were used to calculate an initial estimate of test-retest reliability using Pearson’s R.

**Fig. 1.**
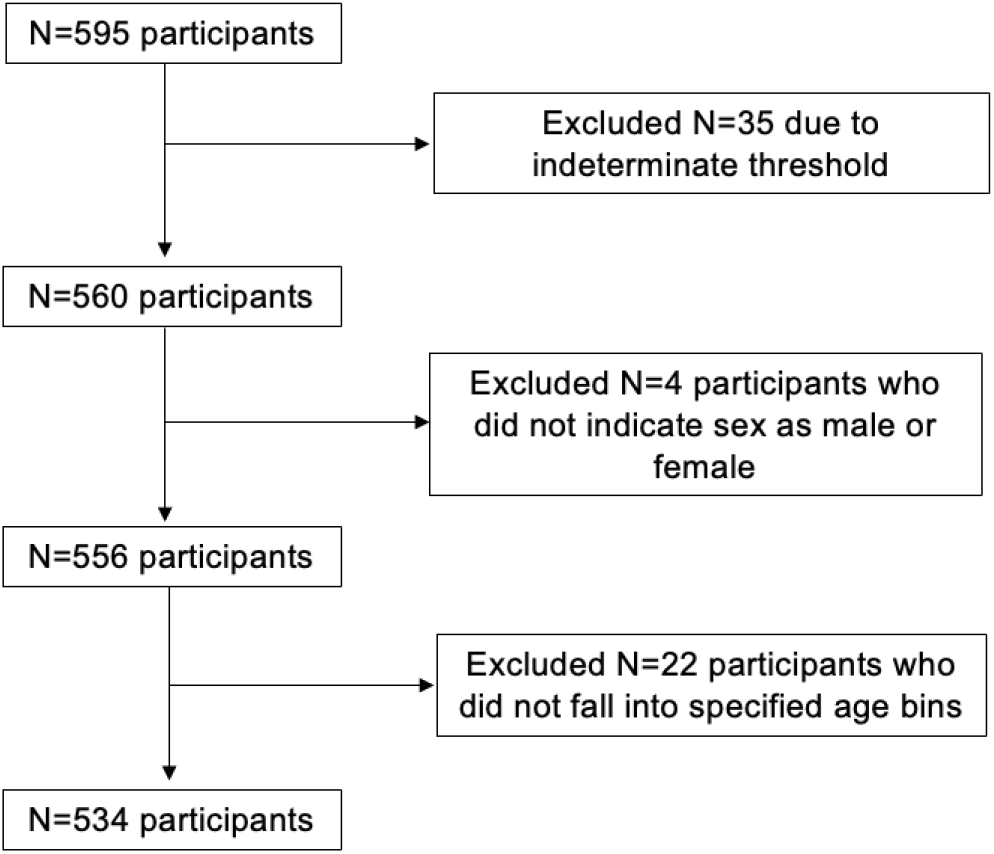
Flow diagram summarizing data cleaning steps resulting in the final participant set used for these analyses (n=534)

Because testing took place at the Twins Day Festival, demographics were enriched in twins and triplets. Of the 534 individuals included in the final data set, 360 were sets of twins (n=180 twin pairs). Of these twin sets, 143 were monozygotic pairs (n=286 individuals) and 37 were dyzogotic pairs (n=74 individuals). Falconer’s formula for broad-sense heritability (H_b_^2^ = 2*(R_mz_ – R_dz_)) was used for a heritability estimate. Uncertainty was calculated by applying the Fisher transformation to Pearson correlations and propagating variance through the calculation (21).

### The ArOMa-T

The ArOMa-T, version 1.P8.1, is composed of a bi-fold card with graphics on the outside; user instructions, along with 17 elliptical Peel-and-Burst™ labels (Scentisphere LLC; Carmel NY) containing an odorant, are on the inside faces of the folded card (Fig. 2). Similar odorant-release technologies have been used in other tests (e.g., (22, 23)). The labels in the ArOMa-T version tested here contain various concentrations of the floral odorant phenylethyl alcohol (PEA) embedded in a proprietary encapsulation matrix. This odorant is used widely in smell testing, including in commercial smell tests marketed by Sensonics International (UPSIT; Haddon Heights, NJ) and Burghart GmbH (Sniffin’ Sticks; Holm, Germany). PEA does not activate the trigeminal nerve at these concentrations, so it is commonly used in odor detection threshold tasks, which do not require any recognition or familiarity of the stimulus name or associated odor. PEA also has abundant normative data in odor detection threshold testing (e.g. (24)), which could facilitate calibration with other assessments. Each ArOMa-T card contains three labels with no odorant, one label each of the lowest and highest odorant concentrations, and two each for the intermediate odorant concentrations. The interval between adjacent concentrations are half-log steps (i.e, 0.5 log_10_ units of concentration, or half an order of magnitude), and the magnitude of the PEA concentration range was chosen to roughly span the range of normal human detection thresholds for PEA (25). The highest concentration contained in the card is arbitrarily denoted as 0 on the log_10_ scale. In this version of the ArOMa-T card (version 1.P8.1), the position of individual PEA concentrations is fixed and unknown to the participants.

**Fig. 2.**
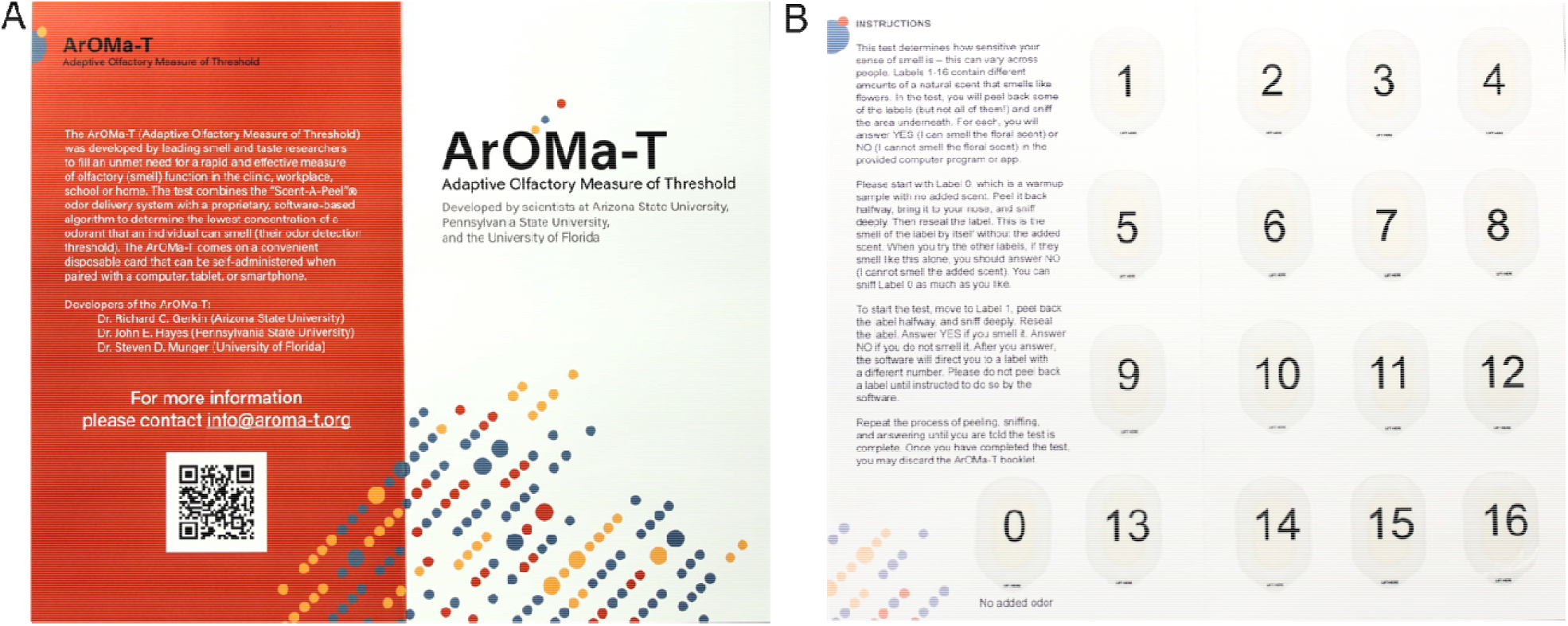
The Adaptive Olfactory Measure of Threshold (ArOMa-T). The test includes a bi-fold card graphics and text on the outside pages **(A)** and with user instruction and 17 peel and burst panels that contain varying amounts of the rose-like/floral odorant phenyl ethyl alcohol (PEA) on the inside pages **(B)**. The unfolded card is approximately 9.5 × 11.15 inches (∼241 mm by ∼286 mm), and the folded card fits easily within a standard 7 in x 10 in envelope for potential delivery via mail. Participants are guided through the task with an accompanying smartphone/tablet app or cloud-based web app that indicates which panel should be peeled and sniffed next. Labels 1-16 contain different PEA concentrations covering a range of ∼3.5 log_10_ units, while label 0 has no added odor as is presented as a reference for the background smell of the label and card. Some labels contain identical concentrations, and some labels are blanks with no PEA added. The blanks and PEA concentrations are distributed across positions in a quasi-random manner so that users cannot infer any information about concentration from position on the card.

The ArOMa-T also employs a unique and novel Bayesian adaptive threshold estimation algorithm (e.g., (26, 27)) to determine which label the user should peel and sniff next (Fig. 3). Before beginning the test, participants are asked to sniff label 0 (which has no added PEA) to familiarize themselves with the background odor of the card. They start the test by sniffing label 1, where two possible responses are considered: “Yes, I can smell it” and “No, I cannot smell it”. All other 15 possibly-odorant-containing labels are then considered by the algorithm as potential choices for the next trial. The specific label selected by the algorithm is the one that is most likely to reduce uncertainty in the running estimate of the detection threshold parameter in the model, weighted by the estimated probability of the “Yes” and “No” responses at the corresponding concentration. This is repeated recursively for all subsequent trials. Because this is very computationally demanding, as the complexity doubles with each additional trial, we have simulated all possible paths in advance to generate a simple lookup-table that is used in the actual application in real time, using a simple web browser application on Apple iPads (9.7in screen; Apple Inc, Cupertino CA). Thus, no internet connection or cloud-based processing was needed to run the algorithm, and test results were stored locally on the iPads. Upon completion, all sensory and demographic data were downloaded for analysis. For this study, only fully de-identified data were accessed.

**Fig. 3.**
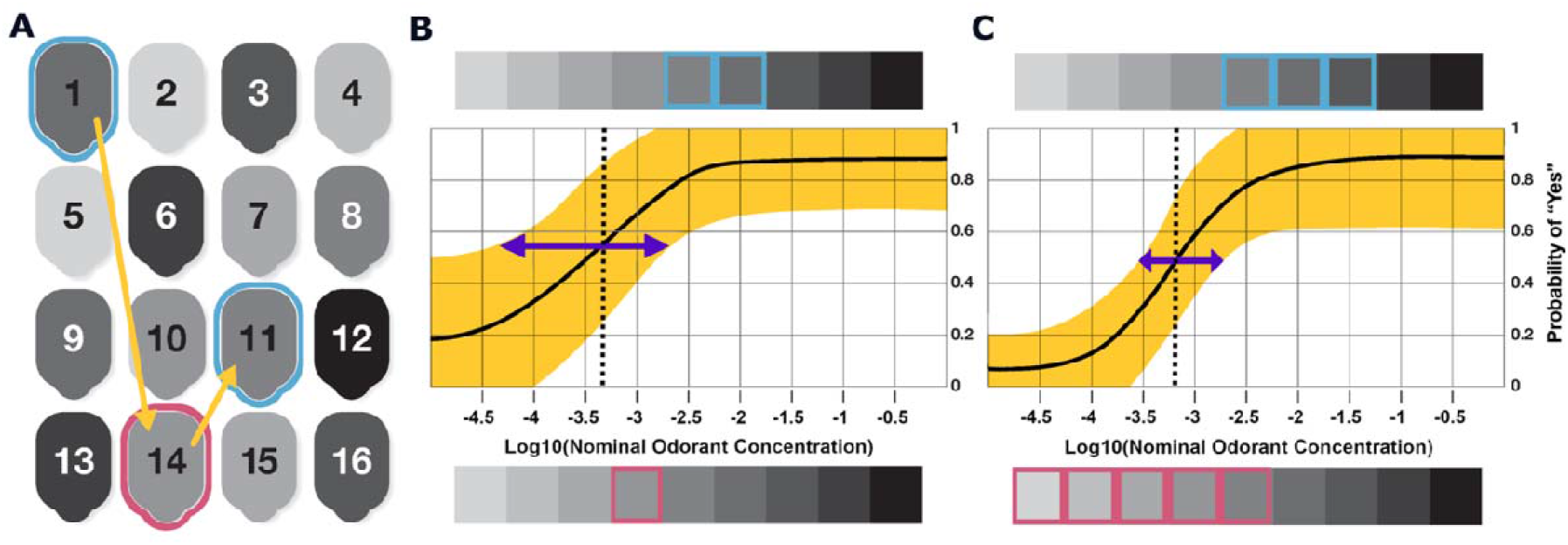
Detection threshold estimation in a single participant using ArOMa-T. **(a)** Schematic of the numbered peel-and-burst labels on the test card, shaded with gray to indicate differing PEA concentration from lowest (light) to highest (dark); in the physical test, users receive no cues about odorant concentration (see Fig. 1). Participants are asked whether they can smell the odor under panel 1 (an intermediate concentration). For a “Yes” response (cyan outline), the app directs them to panel 14; a “No” response to this panel (not shown) would send them to panel 6. A “No” response (magenta outline) for panel 14 directs the subject to panel 11, and so on. The sequence 1, 14, 11 corresponds to one possible path through the first three questions. **(b)** Based on the user’s responses (shown as cyan and magenta rectangles for “Yes” and “No” responses, respectively) and psychometric theory, the algorithm fits a psychometric curve that estimates the probability of a “Yes” response at all concentrations. The solid black line is the point estimate for that curve, and the shaded yellow region is the uncertainty (standard error). The odorant concentration at a “Yes” response probability halfway between the estimated minimum and maximum of that probability is identified as the threshold (vertical dashed line); the uncertainty in this value is indicated with the purple arrow. **(c)** The same user completes an additional five trials in the same test (for a total of eight trials); responses are again shown above and below in the colored rectangles. With this increased number of responses, the estimated curve (and threshold) has shifted, and the uncertainty has been substantially reduced. In our preliminary studies, we used eight trials, but this can be increased or reduced to optimize test accuracy versus time to complete.

### Data collection in volunteers at a festival in Twinsburg Ohio

Due to festival logistics and facilities, as well as COVID-19 pandemic-related safety concerns, all testing occurred outside at ambient temperature. Participants were seated at tables under outdoor canopies on an athletic field. Each participant was provided with an iPad containing the web application to gather demographics and to complete the threshold task. A staff member was available if the participant had questions or technical issues. First, participants entered responses to a few demographic questions, as well as questions on their health history. Next, they self-administered the smell test using the printed ArOMa-T card (Fig. 2) and the preliminary web application described above. Thus, using the iPad and the card, participants were able to self-administer the test.

Each participant was asked to first read the instructions printed directly on the card. They were then instructed to peel and sniff Label 0, which contained no added odor to familiarize themselves with the smell of the card and blank label. Next, the algorithm directed the participant to a label containing an intermediate concentration of PEA (panel 1) and they were asked the binary question: Can you smell the scent (YES/NO). Based on this answer, the algorithm then directed the participant to a label containing a lower concentration of PEA (if the answer was YES) or a higher concentration of PEA (if the answer was NO) (Fig. 3a), where they were again asked to peel the label, sniff, and answer the same YES/NO question. After each question, the algorithm directs the participant to another numbered label based on *all* prior YES/NO responses, which provides a preliminary estimate of detection threshold. Each subsequent response improves this estimate (Fig. 3b). Duplication of some odor concentrations and inclusions of blanks increases the likelihood of faithful responses and allows for criterion bias (e.g., false alarm rate) to be separated from olfactory ability. In the design used in this study, the final detection threshold estimate is determined from a maximum of eight samplings of odorant (and blank) labels, not including label 0. This design allows for a Bayesian adaptive threshold test optimized for speedy self-administration and reporting, and that selects the most informative odorant concentration on each trial.

We used Bayesian inference to estimate the parameters of our psychometric model, applying a weakly informative prior on the detection threshold (log(tau) ∼ *N*(−3, 100)) and on the decision criterion (lambda ∼ *N*(1, 0.5)). Results reported here were robust to a wide range of choices for the mean and variance of the prior, including the lack of a prior (i.e., non-Bayesian inference). Theoretical considerations including biological limits on detection threshold motivated our decision to use a weakly informative prior, as did simulated data checks showing more accurate estimation of model parameters and test/retest reliability in synthetic datasets. This is distinct from logistic regression – such as used in other studies (28-30), which assumes a more rigid probability model for the response.

### Data processing and statistical analysis

Data were analyzed in R using RStudio software (Version 2021.09.0). We determined detection threshold estimates in accordance with the ASTM International (formerly American Society for Testing and Materials) Method E679-19 (“Standard Practice for Determination of Odor and Taste Thresholds by a Forced-Choice Ascending Concentration Series Method of Limits”), with minor modifications as described below. Of the 534 participants tested, 66 participants reported “YES” for all PEA containing labels, and “NO” to all blanks they received; their estimated thresholds were imputed with values slightly below the lowest concentration ArOMa-T cards presents, in accordance with the standard ASTM E679 decision rule. The specific value used here was –4.5 log_10_ units, and these highly sensitive individuals are shown in a box at the left side of figure.

At the other extreme, 23 participants reported “NO” for all concentrations presented; again, following the standard ASTM E679 decision rule, these individuals had their estimated threshold set to a value just above the highest concentration available on the ArOMa-T card. The specific value used here was 0.5 log_10_ units. The seven individuals with threshold values between 0 and 0.5 log_10_ units were successfully fit by the model (i.e., these values reflect their estimated threshold based on a minimal number of YES responses, rather than imputation via an *a priori* decision rule).

### Testing for age & sex differences in individual thresholds determined with the ArOMa-T

A two-way fixed model analysis of variance (ANOVA) was performed to determine if estimated odor detection threshold differed by sex or age group. This was followed by a *post-hoc* comparison with Tukey’s HSD (p< 0.05) to determine where any group differences occurred.

### Exploratory analysis of past COVID-19 status with propensity matched controls

With this convenience sample of participants, this study was not specifically designed or implemented to compare those who experienced a past COVID-19 infection to those who did not; still, the incidence of a prior COVID-19 diagnosis was sufficient to undertake an unplanned exploratory analysis. Participants were not recruited to enrich the sample with COVID-19+ individuals.

Given that only 78 of our participants (∼15%) had a positive self-reported history of COVID-19, we used propensity matching on the participants who indicated they experienced past COVID-19 infection to identify comparable controls without a positive history of COVID-19. Using the MatchIt package (31) in R, participants were matched based on age, sex, and self-identified racial category, thereby generating two equally sized groups of 78 participants each. Due to sex and age group effects in the planned model (Fig. 4), sex and age were included in an exploratory model testing the effect of positive COVID-19 history; due to the smaller sample size (n =156, versus 534 above), age was included in the model as a continuous variable, rather than age group.

**Fig. 4.**
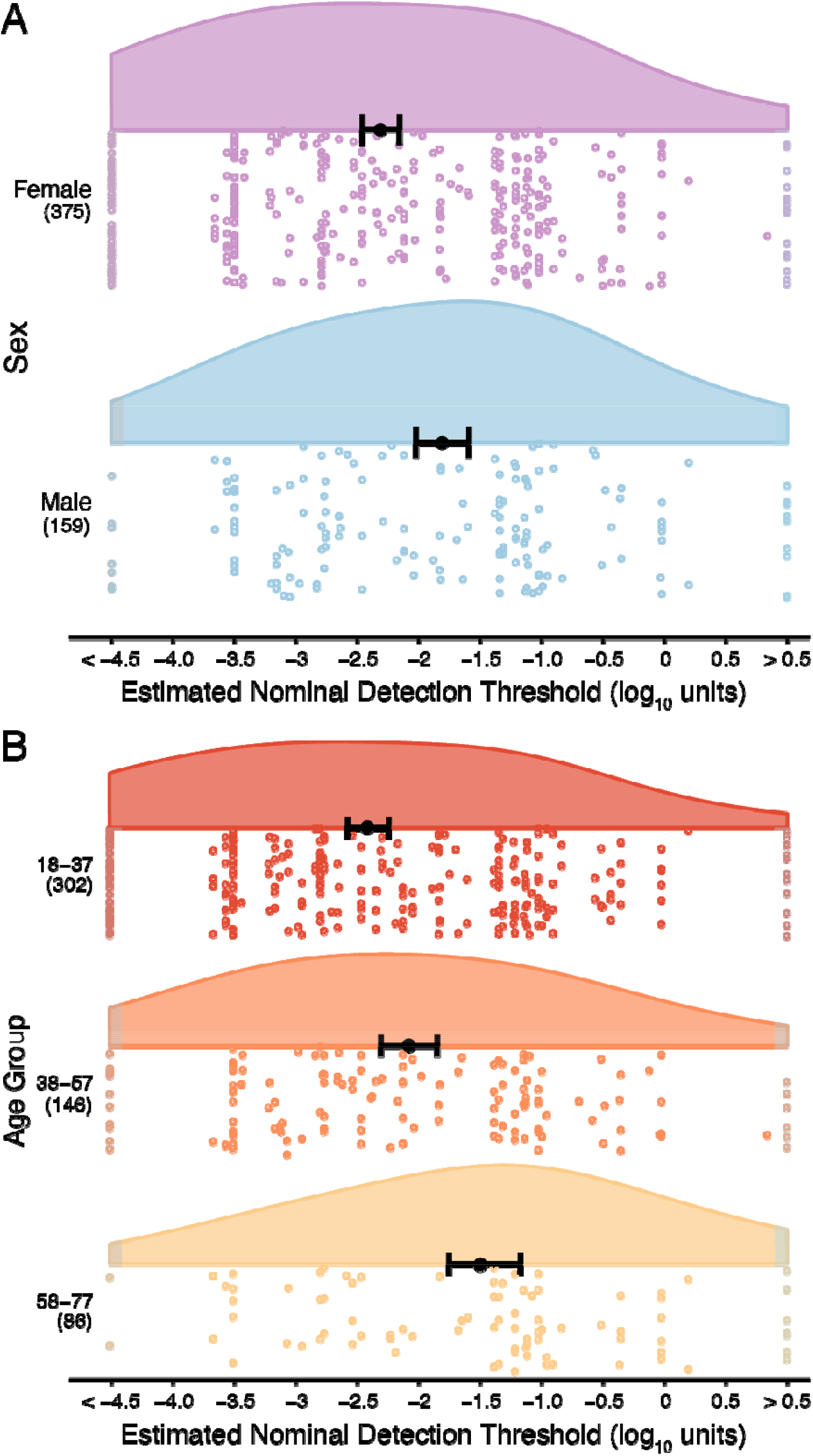
Smell thresholds using the ArOMa-T. **(a)** Raincloud plots showing odor detection thresholds stratified by sex. Open circles represent odor detection thresholds for individual participants; women are shown in purple and men in blue. **(b)** Raincloud plots showing odor detection thresholds stratified by age group, with each age group represented by a different color; again, open circles indicate odor detection thresholds for individual participants. In both panels, the solid black dot is the point estimate of the mean and error bars are 95% confidence intervals of that estimate. Group sample sizes are shown in parentheses on the left side of the plot. Thresholds shown in the boxes on the left and right sides were imputed in accordance with the ASTM E679 rules for extreme values outside the range of the concentrations tested; the light blue box on the right highlights functionally anosmic individuals with thresholds above the range tested here, while the gray box on the left indicates highly sensitive normosmic individuals who responded YES to all odorant concentrations. The detection threshold (X-axis) is expressed in the base-10 logarithm of the nominal concentrations.

A three-way fixed ANOVA was used to test whether estimated odor detection threshold differed by past COVID-19 status, adjusting for sex and age (as a continuous variable) followed by a post-hoc comparison using Tukey’s HSD (p<0.05). In parallel, a two sample Kolmogorov-Smirnov (KS) test was also performed to test for differences in distribution shape.

## 3. Results

### Administration of the ArOMa-T

In a non-laboratory setting with an age diverse set of participants, the mean time to complete the ArOMa-T was 2.8 ± 0.9 min, inclusive of time needed to read the instructions. Individual trials – i.e., sniffing a label and answering a single YES/NO question – took an average of 11.6 ± 2.8 sec. The false alarm rate for the ArOMa-T (answering “YES” to the first blank presented) was 7.5%, while the rate of answering incorrectly to *two blanks* was 2.6%. A subset of participants (n=97) retook ArOMa-T the following day. For this subgroup, we found a Pearson’s R of 0.61, comparable to the published test-retest reliability of 0.58 for another rapid olfactory measure, the NIH Toolbox Odor Identification Test (Toolbox OIT) (32).

### Effect of Sex and Age on Thresholds

There was a main effect of sex on estimated odor threshold measured with the ArOMa-T [F(1,528) = 15.97; p<0.0001]. Female participants showed a detection threshold that was ∼0.50 log_10_ units (∼3.2x) lower (i.e., more sensitive) than males: − 2.31 versus −1.81, respectively Fig. 4a. There was a greater percentage of females (15.5%) than males (5.0%) with the lowest measurable detection threshold (−4.5 log_10_ units).

There was a main effect of age-group on estimated odor threshold [F(2,528) = 15.32; p<0.0001; Fig. 4b]. In Tukey’s HSD (p<0.05), the youngest participants (18-37 years) had a lower mean threshold estimate compared to participants in oldest group (58-77 years). The size of this difference was ∼0.94 log_10_ units (a ∼8.7x difference in concentration). The participants in the second age bin (38-57 years) had a lower mean threshold estimate compared to participants in oldest group (58-77 years). The size of this difference was ∼0.61 log_10_ units (a ∼4.1x difference in concentration). Finally, there was no evidence of an interaction between sex and age-group [F(2,528) = 0.32; p=0.73].

### Propensity matching and past COVID-19 status

The ANOVA model testing for an effect of positive COVID-19 history on smell threshold did not find an association between COVID-19 history and odor detection threshold [F(1,152)=0.11; p=0.74]. Specifically, the mean threshold estimate of participants without a past history of COVID-19 was −1.96 ± 1.46, compared to −1.88 ± 1.30 for participants who self-reported a prior COVID-19 infection, a nominal difference of ∼0.08 log units (∼1.2x difference). Likewise, we found no evidence for a difference in the threshold distributions between participants who previously had COVID-19 and those who did not (KS test statistic = 0.09; p=0.91). Collectively, these exploratory analyses provide no evidence that odor thresholds are elevated in a convenience sample of individuals who have previously recovered from acute COVID-19. However, this finding is only tentative and should be confirmed in larger samples with specific study recruitment intended for such comparisons.

### Heritability of Detection Thresholds measured with the ArOMa-T

The large number of both monozygotic and dizygotic twins in our sample allowed us to conduct a preliminary estimate of heritability for detection threshold. Dizygotic twins exhibited weak correlation in estimated detection thresholds (R=0.19 ± 0.16), while monozygotic twins showed a stronger correlation (R=0.46 ± 0.07). Using Falconer’s formula (see Methods), we estimate a broad-sense heritability of H^2^ = 0.55 ± 0.36.

## 4. Discussion

The primary goal of this study was to assess the ability of a novel smell test, the ArOMa-T, and to determine an odor detection threshold in individuals in a non-clinical setting. While exact odor detection threshold estimates depend on the specific psychophysical method used to operationally measure the threshold (20), threshold-based assessments have substantial advantages over other measures of olfactory function as they avoid issues of prior familiarity, memory recall, and naming ability. However, the administration of other threshold-based methods can be tedious and slow, and thus are often avoided in lieu of other methods when time is at a premium (i.e., clinical visits).

### Administration of the ArOMa-T

Here, we found the ArOMa-T is a fast, easy to use, field-deployable test. Over 500 participants were able to complete the test despite its being administered in an outdoor, festival setting. The median time to complete an individual ArOMa-T – under three minutes – compares favorably to commercially available smell tests like the UPSIT and Sniffin’ Sticks, which can take eight or more minutes to complete. When participants were presented with two blanks (versus one), the false alarm rate – the fraction of participants who responded YES to all blanks – was dramatically reduced from 7.5% to 2.6%. In future studies or in clinical use, it will be trivial to adjust the algorithm to require multiple blank trials for every participant. Further, with a Pearson’s R of 0.61, the test-retest reliability of ArOMa-T is comparable to other validated self-administered rapid smell tests, such as the NIH Toolbox Odor Identification Test (Toolbox OIT), which has a test-retest reliability of 0.58 (32). The Pearson’s R observed here for the ArOMa-T suggests this test is both reliable (i.e., it provides an accurate representation of a participant’s performance across testing sessions) and internally valid.

### Effects of sex on olfactory threshold detection

This study recapitulated well-known sex differences in odor thresholds that show females have higher olfactory sensitivity when compared to males (Fig 4) (17, 18, 33). Females have consistently been shown to be more sensitive than males for many odorants, including 1-butanol, 1-hexanol, and 1-octanol (34) (35). While there is no definitive explanation for the common observation that females show superior olfactory performance than males, hormone interactions of sex hormones with the olfactory system have been suggested (35-37). Separately, females may have higher ability to pick up odors from a multitude of external stimuli, known as odor awareness (38). Last, it has been suggested previously that the rate of olfactory decline is greater among males (39) and that males may be more prone to harmful occupational exposure to toxic compounds that can damage the olfactory system (40). Regardless of the underlying reasons for these sex differences, however, it is clear that the ArOMa-T has sufficient sensitivity to discern expected sex differences.

### Effects of age on olfactory threshold detection

We also found that average odor detection thresholds increased with age (Fig. 4), similar to findings from previous studies (17, 18). Notably, younger participants were more likely to exhibit the lowest odor detection thresholds. The higher average odor detection thresholds seen in the oldest participants for this study is consistent with previous studies that found about half of the U.S. population between the ages of 65-80, and about three-quarters over the age of 80, experience smell loss (e.g., (41)). The relationship between age and olfactory decline has been seen when only odor identification ability was assessed (42), or when odor identification, discrimination and detection threshold were all tested (17). Indeed, odor identification and threshold tests are both sensitive to age-related smell loss (16, 19), consistent with present data gathered with the ArOMa-T.

### Effects of past COVID-19 status on olfactory threshold detection

Recent studies have explored olfactory detection thresholds as a measure of smell loss due to current or prior SARS-CoV-2 infection (e.g., (43, 44)). One study found that odor detection threshold scores were more affected than discrimination and identification scores in COVID-19 patients tested using Sniffin’ Sticks odor wands at least two weeks after symptom onset (43). Elsewhere, it was reported that the majority of COVID-19 patients had impaired olfactory thresholds when tested with the Connecticut Chemosensory Clinical Research Center orthonasal olfaction test approximately two weeks after a positive SARS-CoV-2 PCR test (44). Further, it has been reported that olfactory threshold scores from Sniffin’ Sticks were more affected than scores for odor identification and discrimination in hospitalized patients with COVID-19 tested ∼1 month after diagnosis (45). Collectively, these and other reports suggest detection threshold may be particularly susceptible to COVID-19; if confirmed, this might suggest some individuals with COVID-19 experience hyposmia that is missed with an odor identification task.

The relationship between subjective assessment of smell and controlled psychophysical testing remains highly contentious (see (46)). Self-report can be subject to recall bias, and many with measurable loss may be unaware of this loss (41, 47). On the other hand, measures of subjective loss or dysfunction may better capture quality of life issues, including dietary intake (48). Notably, qualitative disorders like parosmia and phantosmia can only be assessed via patient history and self-report, as no objective tests exist for these conditions (3). Our results conflict somewhat with some prior work, as we saw no convincing evidence that thresholds were elevated in those who had recovered from COVID-19. However, this was a small convenience sample of individuals without active COVID-19, and we have no estimate of the elapsed time between illness and olfactory testing. Additional work in larger cohorts with recruitment stratified by current COVID-19 status and/or past history is warranted to resolve these questions. The ArOMa-T may be especially well-suited for such study designs, given that it is mailable, rapid, and suitable for field use.

## Limitations

Several limitations should be noted. Foremost, the sample was predominantly non-Hispanic white, so results may not generalize to other demographic groups. Also, the test setting could have biased recruitment of older individuals towards those in exceptionally good health – i.e., a ‘healthy worker effect’ (49). This may partially explain why some participants in the 58-77 age category show the same or a lower olfactory detection threshold as those in the 18-37 age category. Thus, we cannot make sweeping generalizations about aging. Further, our heritability estimate is highly uncertain, as the sample size (of twin pairs) is too small. While PEA is commonly used in olfactory testing in part because specific anosmias for this compound are rare, we cannot exclude that performance on this test may vary as a function of individual genetics. The moderate heritability observed here could reflect numerous causes of differences in overall olfactory ability including differences in specific anosmias, nasal patency, or myriad other factors that are shared by monozygotic siblings. Additional work in larger and more deliberately stratified samples will be necessary to resolve these questions. Finally, while our test is deliberately designed to separate response bias from underlying ability, some biases (such as malingering) would be difficult to distinguish from anosmia without additional assessments.

## Conclusions

We found that the ArOMa-T can be used as a rapid olfactory screening test to capture variation in detection thresholds. We observed differences in estimated detection thresholds between sex and age groups, highlighting the ability of the ArOMa-T to reproduce expected population level findings, as well as demonstrating its ability serve as a portable and rapid smell test. This is highly advantageous compared to other olfactory tests that take longer to complete, have portability limitations, or must be administered by a trained individual. In a field-based convenience sample, no evidence was found to suggest detection thresholds differ between participants who report a history of COVID-19 and matched controls who did not. However, these tentative null results require confirmation in a study specifically designed to explore this question. Collectively, our results suggest the ArOMa-T is able to reproduce sex and age effects previously observed with more time-intensive testing methods.

## Data Availability

All fully de-identified data are available upon reasonable request to the authors.

## Acknowledgments

The authors warmly thank our participants for their time and critical contribution to this work. We also thank Samantha Fadool for her design of the artwork for the ArOMa-T version used here.

